# High Variability of Body Mass Index Independently Associated with Incident Heart Failure

**DOI:** 10.1101/2023.03.30.23287990

**Authors:** Chang Liu, Yiyun Chiang, Qin Hui, Jin J. Zhou, Peter W.F. Wilson, Jacob Joseph, Yan V. Sun

**Affiliations:** Department of Epidemiology, Emory University Rollins School of Public Health, Atlanta, Georgia, USA; Department of Medicine and Biostatistics, University of California, Los Angeles, California, USA; Atlanta VA Health Care System, Decatur, GA, USA; Department of Medicine, Emory University School of Medicine, Atlanta, GA, USA; VA Providence Healthcare System, Providence, Rhode Island, USA; Warren Alpert Medical School, Brown University, Providence, Rhode Island, USA

**Keywords:** Body Mass Index, BMI, Heart Failure, Heart Failure Incidence

## Abstract

**Background:** Heart failure (HF) is a serious condition with increasing prevalence, high morbidity, and increased mortality. Obesity is an established risk factor for cardiovascular diseases, including HF. Fluctuation in body mass index (BMI) has shown a higher risk of cardiovascular outcomes. We investigated the association between BMI variability and incident HF.

**Methods:** In the UK Biobank, we established a prospective cohort after excluding participants with prevalent HF or cancer at enrollment. A total of 99,368 White (British, Irish, and any other white background) participants with ≥ 3 BMI measures during > 2 years preceding enrollment were included, with a median follow-up of 12.5 years. The within-participant variability of BMI was evaluated using standardized standard deviation (SD) and coefficient of variation (CV). The association of BMI variability with incident HF was assessed using Fine and Gray’s competing risk model, and adjusted for age, sex, smoking history, alcohol consumption, diabetes, hypertension, history of heart attack, stroke, atrial fibrillation, lipids, estimated glomerular filtration rate and mean BMI per individual.

**Results:** In the fully adjusted model, higher BMI variability measured in both SD and CV were significantly associated with higher risk in HF incidence (SD: Hazard Ratio [HR] 1.05, 95% Confidence Interval [CI] 1.02 - 1.07, p = 0.0002; CV: HR 1.06, 95% CI 1.04 - 1.09, p < 0.0001).

**Conclusions:** Longitudinal health records capture BMI fluctuation, which independently predicts HF incidence. Integration of long-term BMI and other routinely measured health factors may improve risk prediction of HF and other cardiovascular outcomes.

## Introduction

Heart failure (HF) is a disease of high public health burden. It affects an estimated total of 64 million HF patients worldwide ^1^, including 6 million in the US ^2^. The prevalence of HF is projected to exceed 8 million adults by the year 2030 ^3^. HF prevalence is similar among both males and females, however a higher incidence has been observed among males and a longer survival was observed among females ^4^. Many factors play critical roles in HF etiology, such as obesity ^5^, aging ^6,7^, diabetes ^8^, hypertension ^9^. Their potential causal effects were supported by recent Mendelian Randomization studies ^10,11^.

Further, as one of the leading risk factors for HF, obesity is associated with a high population attributable fraction of up to 21% in HF cases ^12,13^. Fluctuation in body mass index (BMI) has a negative impact on health ^14-17^, such as associations with higher risk in coronary heart disease, coronary heart disease mortality and all-cause mortality ^14-17^. While the association between BMI variability and HF was identified among patients with type 2 diabetes ^18^ with possible explanation of metabolic syndrome development, such association has not been established among the general population. Herein, we assessed the hypothesis that BMI variability is associated with incident HF among a large cohort free of prevalent HF and cancer, independent of mean BMI.

## Methods

### Study Population

This study was carried out using the data from the UK Biobank, a large prospective cohort study with participants aged 40-69 enrolled in 2006-2010 ^19^. Longitudinal measures of BMI were obtained from the baseline measure and linkage to primary care data ^19^, which includes approximately 42% of the UK Biobank participants having ≥ 1 BMI records identified from the primary care data. Inclusion criteria for this current study include: White (British, Irish, and any other white background) participants free of HF at enrollment; participants with ≥ 3 BMI measures from primary care records within a time window of > 2 years before and at enrollment, to assess the BMI variability. HF onset was ascertained based on linkage to Hospital Episode Statistics (HES) data, using International Classification of Diseases (ICD) -10 codes I11.0, I13.0, I13.2, I25.5, I42.0, I42.5, I42.8, I42.9, I50.0, I50.1, I50.9, and self-reported medical conditions during enrollment interview ^19^. The earliest date of diagnosis was compared with the enrollment date, and the HF prevalent cases before or at enrollment were excluded, while the HF incident cases after enrollment were treated as outcomes of interest. To avoid possible weight loss due to malignancy, we excluded participants with cancer diagnosed before or at enrollment using the national cancer registries linked with the UK Biobank ^19^. A total of 99,368 participants with a median follow-up of 12.5 years (interquartile range 12.0-13.3 years) were included in this study based on aforementioned inclusion and exclusion criteria. Time to incident HF was defined as time compared with enrollment date for one of the following: incident HF diagnosis after enrollment, loss to follow-up, or end of follow-up. Information concerning ejection fraction for the HF incident cases was not available.

### Statistical Analysis

For the overall cohort and separately by sex, characteristics were summarized as mean ± standard deviation or median and interquartile range for continuous variables, and count (percent) for categorical variables. Mann-Whitney U test for continuous variables, and Chi-squared test for categorical variables were used to compare between incident heart failure vs. no incident heart failure where appropriate. The within-participant variability of BMI was calculated using standard deviation (SD) and coefficient of variation (CV, SD divided by within-participant mean BMI) across the multiple measurements. The SD and CV were mean centered and scaled by standard deviation for downstream analyses, for an interpretation based on a standardized scale.

Kaplan-Meier curves coupled with log-rank tests were used to visualize the difference in HF incidence between the groups with highest vs. lowest 20% quantile of BMI variability measured in both SD and CV. The association of BMI variability with incident HF was assessed using Fine and Gray’s competing risk model, treating death events as competing risk events ^20^. Two models were assessed: Model 1 adjusted for age, sex, smoking history (current or previous smoking vs. never smoking), frequent alcohol consumption (≥ 3 times per week vs. less frequent), self-reported health conditions including diabetes, hypertension, heart attack history, stroke history, and ICD - 10 (I44.0, I44.1, I44.2, I44.3, I44.5, I46.0, I46.1, I46.9, I47.0, I47.2, I48, I48.1, I48.2, I48.3, I48.4, I48.9, I49.0, I49.5) defined atrial fibrillation history based on HES data, high density lipoprotein cholesterol, low density lipoprotein, total cholesterol, triglycerides, and estimated glomerular filtration rate estimated using the 2021 CKD-EPI equation ^21^; Model 2 additionally adjusted for within-participant mean BMI across longitudinal measures. Using Model 2 and excluding risk factor of stratification, we further explored the associations across risk factor strata: sex, age (<60 years or ≥ 60 years), diabetes, hypertension, and BMI at enrollment categorized as underweight (BMI < 18.5 kg/m^2^), normal weight (BMI ≥ 18.5, < 25 kg/m^2^), overweight (BMI ≥ 25, < 30 kg/m^2^) and obese (BMI ≥ 30 kg/m^2^). The interaction between BMI variability and these risk factors was also tested using Model 2.

Analyses were performed using R version 4.0.2 (R Foundation for Statistical Computing, Vienna, Austria) and SAS statistical software version 9.4 (SAS Institute Inc, Cary, NC).

## Results

The cohort of 99,368 participants was 57% female, with mean age 57.5 (SD 7.8) years. A total of 3,406 incident HF cases were observed during the follow-up, **Table 1**. Participants who experienced incident heart failure had a higher level of BMI variability in SD compared to participants who did not (SD: 1.68 vs. 1.53, p < 0.001), however not in CV (0.0562 vs. 0.0560, p = 0.137). Participants who experienced incident heart failure also had a lower proportion of female (35.6% vs. 57.8%, p < 0.001), higher proportions in lifestyle risk factors and prevalence of cardiovascular comorbidities, medication use, higher mean BMI and BMI at enrollment, higher levels in lipid profile except for high- and low-density lipoprotein cholesterol, **Table 1**.

**Table 1.** Cohort Characteristics.

| **Variables** | **Overall Cohort**<br>**N=99,368** | **Incident Heart**<br>**Failure**<br>**N=3,406**<br>**(3.4%)** | **No Incident Heart**<br>**Failure**<br>**N=95,962**<br>**(96.6%)** | **P\*** |
| --- | --- | --- | --- | --- |
| Female (%) | 56,655 (57.0%) | 1,213 (35.6%) | 55,442 (57.8%) | **<0.001** |
| Age at enrollment, years | 57.5 ± 7.8 | 62.5 ± 5.8 | 57.4 ± 7.8 | **<0.001** |
| Smoking history (%) | 45,430 (45.7%) | 2,053 (60.3%) | 43,377 (45.2%) | **<0.001** |
| Frequent alcohol consumption (%) | 42,611 (42.9%) | 1,356 (39.9%) | 41,255 (43.0%) | **<0.001** |
| Diabetes (%) | 7,098 (7.1%) | 728 (21.4%) | 6,370 (6.7%) | **<0.001** |
| Hypertension (%) | 33,249 (33.5%) | 1,951 (57.4%) | 31,298 (32.7%) | **<0.001** |
| Heart attack history (%) | 2,986 (3.0%) | 571 (16.8%) | 2,415 (2.5%) | **<0.001** |
| Stroke history (%) | 1,859 (1.9%) | 217 (6.4%) | 1,642 (1.7%) | **<0.001** |
| Atrial fibrillation history (%) | 1,689 (1.7%) | 328 (9.6%) | 1,361 (1.4%) | **<0.001** |
| BMI at enrollment, kg/m<sup>2</sup> | 28.0 ± 5.1 | 30.55 ± 6.1 | 27.88 ± 5.0 | **<0.001** |
| Within-participant mean BMI<br>(across longitudinal measures), kg/m<sup>2</sup> | 27.0 ± 4.7 | 29.4 ± 5.6 | 26.9 ± 4.6 | **<0.001** |
| HDL-C, mg/dL | 55.8 ± 14.8 | 49.9 ± 14.1 | 56.0 ± 14.8 | **<0.001** |
| LDL-C, mg/dL | 136.0 ± 34.3 | 122.2 ± 35.7 | 136.6 ± 34.1 | **<0.001** |
| Total cholesterol, mg/dL | 218.0 ± 45.2 | 227.0 ± 43.9 | 207.1 ± 44.4 | **<0.001** |
| Triglycerides, mg/dL (median, IQR) | 134.3 [95.0, 193.4] | 151.7 [106.7, 214.7] | 133.7 [94.6, 192.6] | **<0.001** |
| eGFR, mL/min/1.73 m<sup>2</sup> | 93.9 ± 13.1 | 87.6 ± 16.4 | 94.1 ± 13.0 | **<0.001** |
| Cholesterol lowering medication (%) | 22,611 (22.9%) | 1,774 (52.4%) | 20,837 (21.8%) | **<0.001** |
| Blood pressure medication (%) | 12,308 (12.5%) | 541 (16.0%) | 11,767 (12.3%) | **<0.001** |
| **BMI variability** |  |  |  |  |
| Standard deviation (SD) | 1.54 ± 1.26 | 1.68 ± 1.24 | 1.53 ± 1.26 | **<0.001** |
| Coefficient of variation (CV) | 0.0560 ± 0.0393 | 0.0562 ± 0.0396 | 0.0560 ± 0.0394 | 0.137 |

Mean ± standard deviation shown for continuous variables unless stated otherwise; count (percent) shown for categorical variables.
BMI, body mass index; HDL-C, high density lipoprotein cholesterol; LDL-C, low density lipoprotein cholesterol; eGFR, estimated glomerular filtration rate.
\*P value for comparing incident heart failure vs. no incident heart failure groups.

Participants with highest quintile of BMI variability measured in both SD (p < 0.0001) and CV (p = 0.011) showed a higher incidence of HF compared to the lowest quintile (**Figure 1, Figure 2 and Graphical Abstract)**. In Model 1, the effect per positive SD in BMI variability, measured in both SD and CV, was associated with greater risk in HF incidence (SD: Hazard Ratio [HR] 1.12, 95% Confidence Interval [CI] 1.1-1.14, p < 0.0001; CV: HR 1.11, 95% CI 1.08-1.14, p < 0.0001). After additionally adjusting for within-participant mean BMI in Model 2, the associations were reduced but remained statistically significant (SD: HR 1.05, 95% CI 1.02-1.07, p = 0.0002; CV: HR 1.06, 95% CI 1.04-1.09, p < 0.0001) (**Table 2 and Figure 3)**. In Model 2, the within-participant mean BMI across longitudinal measures was associated with higher risk of incident HF (**Supplementary Table 1)**.

**Figure 1:**
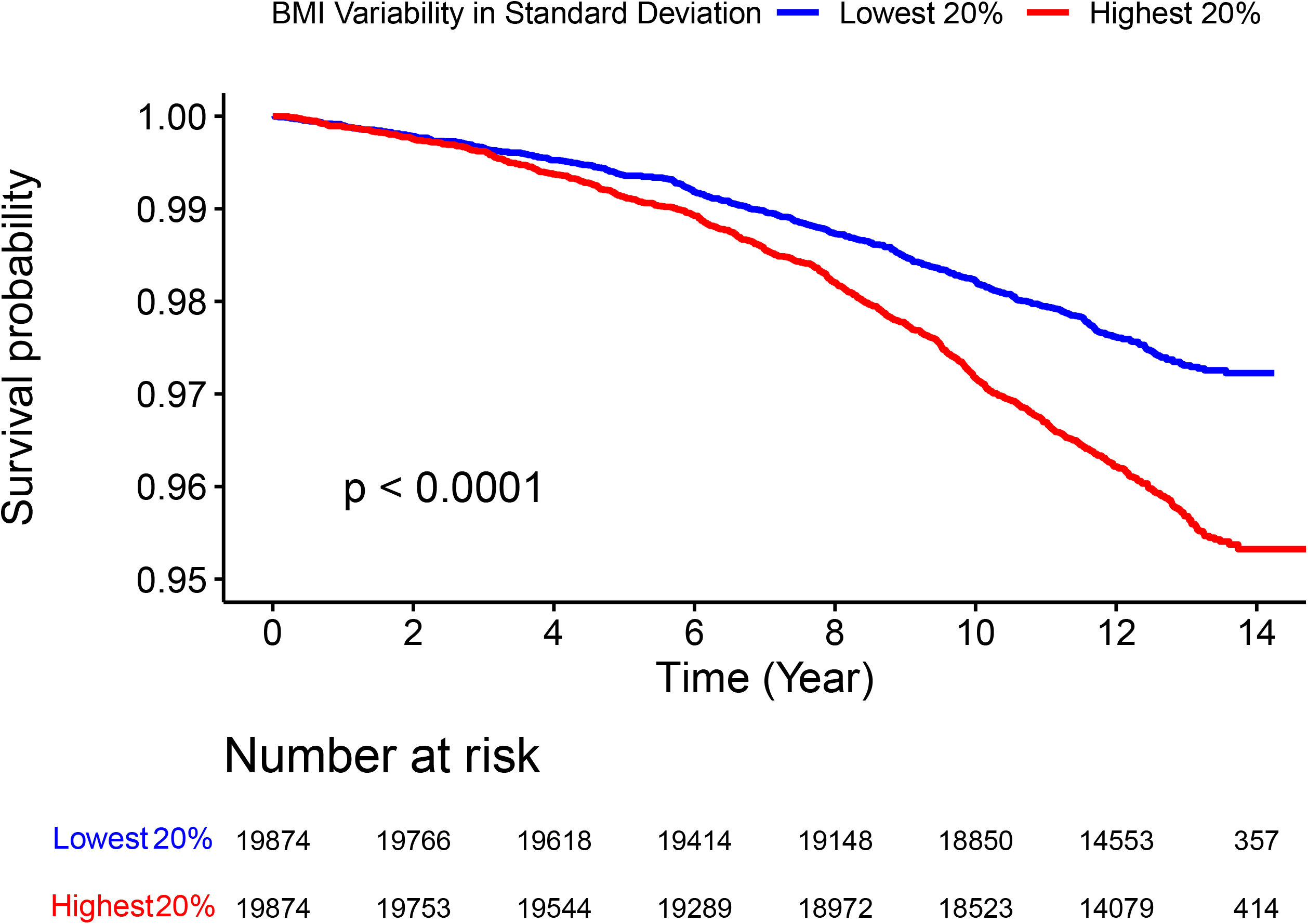
Kaplan-Meier Curve Comparing HF Incidence Between Highest (Red) and Lowest (Blue) 20% Quantile of BMI Variability Measured in Standard Deviation. Log-rank test p value indicates statistically significant difference in HF incidence between the groups.

**Figure 2:**
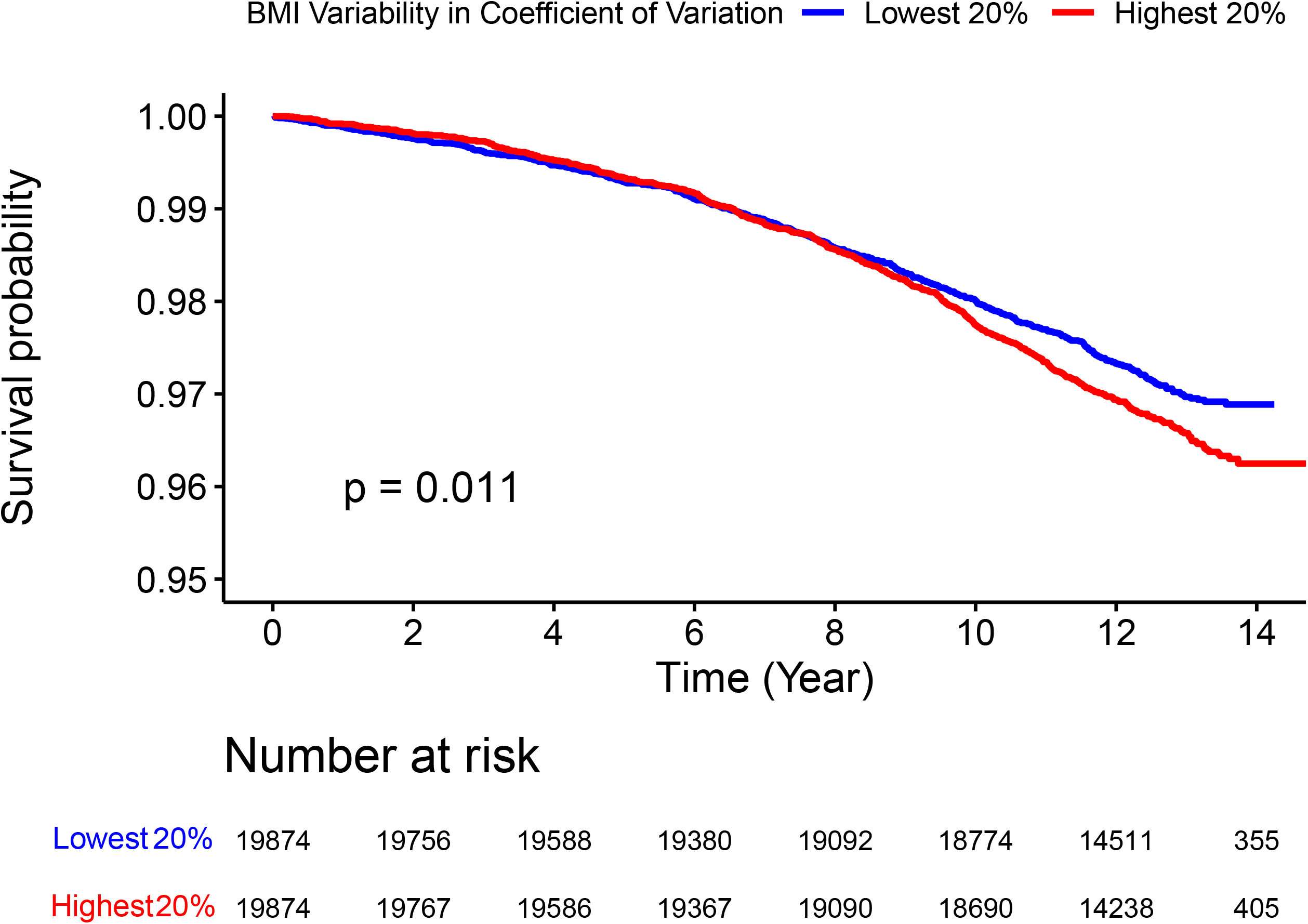
Kaplan-Meier Curve Comparing HF Incidence Between Highest (Red) and Lowest (Blue) 20% Quantile of BMI Variability Measured in Coefficient of Variation. Log-rank test p value indicates statistically significant difference in HF incidence between the groups.

**Table 2:** Association Between BMI Variability (Measured in SD and CV, per standard deviation) and Incident Heart Failure.

| Cohort | SD |  | CV |  |
| --- | --- | --- | --- | --- |
|  | HR (95% CI), P |  | HR (95% CI), P |  |
|  | Model 1 | Model 2 | Model 1 | Model 2 |
| Overall | 1.12 (1.1, 1.14),<br>p<0.0001 | 1.05 (1.02, 1.07),<br>p=0.0002 | 1.11 (1.08, 1.14),<br>p<0.0001 | 1.06 (1.04, 1.09),<br>p<0.0001 |
| Female | 1.15 (1.11, 1.19),<br>p<0.0001 | 1.07 (1.03, 1.11),<br>p=0.0009 | 1.14 (1.09, 1.19),<br>p<0.0001 | 1.09 (1.04, 1.14),<br>p<0.0001 |
| Male | 1.10 (1.08, 1.13),<br>p<0.0001 | 1.03 (1, 1.06),<br>p=0.0674 | 1.09 (1.05, 1.13),<br>p<0.0001 | 1.04 (1.01, 1.08),<br>p=0.0178 |
| Age < 60 years | 1.15 (1.11, 1.19),<br>p<0.0001 | 1.07 (1.02, 1.12),<br>p=0.0024 | 1.14 (1.09, 1.2),<br>p<0.0001 | 1.09 (1.04, 1.15),<br>p=0.0004 |
| Age ≥ 60 years | 1.11 (1.09, 1.14),<br>p<0.0001 | 1.04 (1.01, 1.07),<br>p=0.0076 | 1.1 (1.07, 1.14),<br>p<0.0001 | 1.05 (1.02, 1.09),<br>p=0.0006 |
| No Diabetes | 1.11 (1.09, 1.14),<br>p<0.0001 | 1.04 (1.01, 1.07),<br>p0.0023 | 1.11 (1.08, 1.14),<br>p<0.0001 | 1.06 (1.03, 1.09),<br>p<0.0001 |
| Diabetes | 1.17 (1.11, 1.23),<br>p<0.0001 | 1.08 (1.02, 1.16),<br>p=0.0119 | 1.15 (1.08, 1.22),<br>p<0.0001 | 1.10 (1.03, 1.18),<br>p=0.0053 |
| No Hypertension | 1.11 (1.09, 1.14),<br>p<0.0001 | 1.05 (1.01, 1.08),<br>p=0.0037 | 1.11 (1.08, 1.15),<br>p<0.0001 | 1.06 (1.03, 1.1),<br>p=0.0003 |
| Hypertension | 1.13 (1.1, 1.17),<br>p<0.0001 | 1.05 (1.01, 1.09),<br>p=0.0157 | 1.11 (1.07, 1.16),<br>p<0.0001 | 1.06 (1.02, 1.11),<br>p=0.003 |
| **Normal Weight**<br>(Enrollment BMI $\geq 18.5$, $< 25$ kg/m<sup>2</sup>) | **1.07 (1.01, 1.13),**<br>**p=0.03** | **1.09 (1.02, 1.18),**<br>**p=0.016** | **1.08 (1.02, 1.15),**<br>**p=0.0078** | **1.10 (1.03, 1.18),**<br>**p=0.0068** |
| **Overweight**<br>(Enrollment BMI $\geq 25$, $< 30$ kg/m<sup>2</sup>) | **1.07 (1.03, 1.11),**<br>**p=0.0008** | **1.06 (1.01, 1.11),**<br>**p=0.0118** | **1.08 (1.03, 1.13),**<br>**p=0.0007** | **1.08 (1.02, 1.13),**<br>**p=0.0029** |
| **Obese**<br>(Enrollment BMI $\geq 30$ kg/m<sup>2</sup>) | **1.13 (1.08, 1.17),**<br>**p<0.0001** | **1.06 (1.02, 1.11),**<br>**p=0.0039** | **1.09 (1.04, 1.15),**<br>**p=0.0004** | **1.08 (1.03, 1.14),**<br>**p=0.0011** |

**Figure 3:**
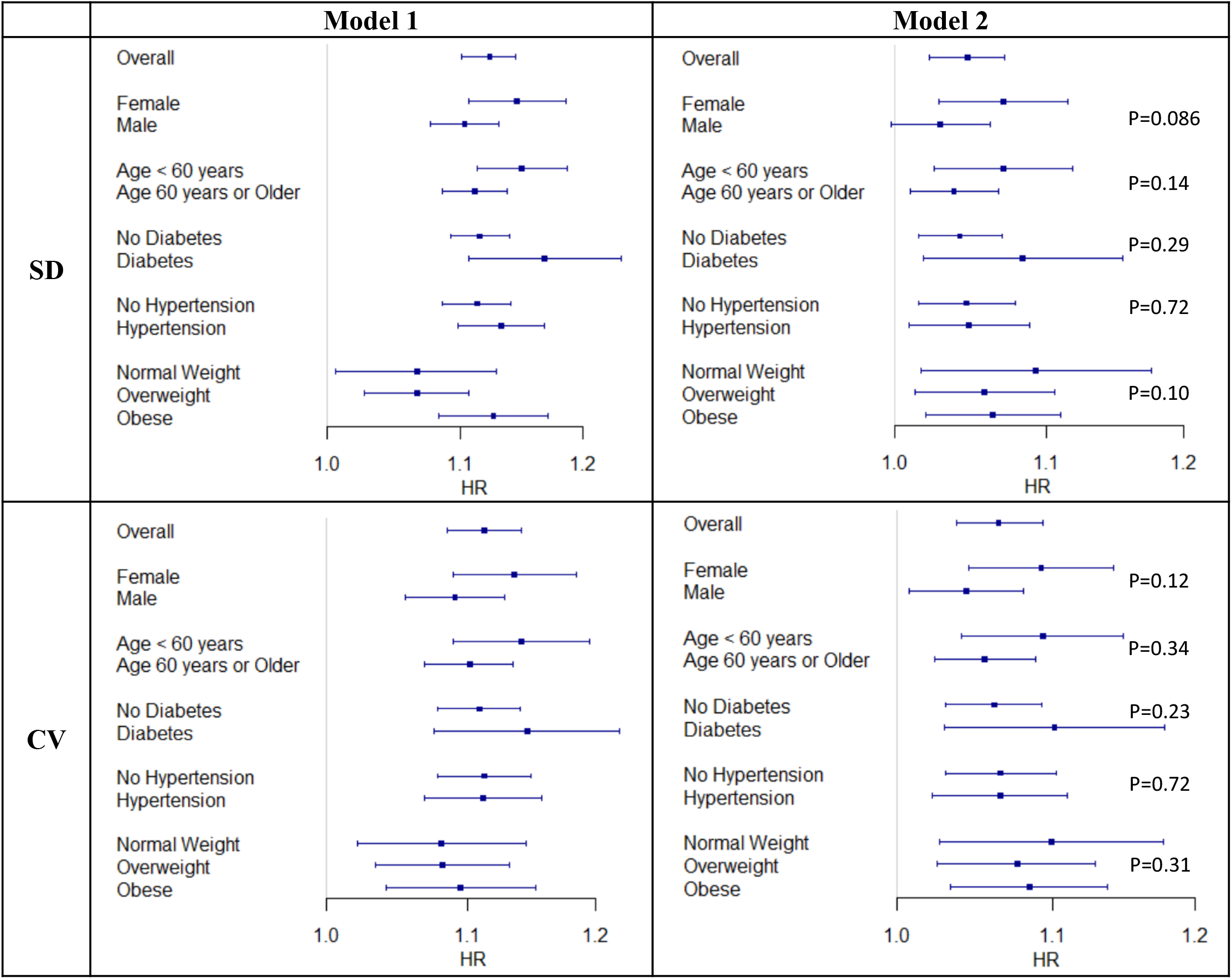
Association Between BMI Variability (Measured in SD and CV, per standard deviation) and Incident Heart Failure. Model 1 adjusted for age, sex, smoking history (current or previous smoking vs. never smoking), frequent alcohol consumption (≥ 3 times per week vs. less frequent), diabetes, hypertension, heart attack history, stroke history, atrial fibrillation history, high density lipoprotein cholesterol, low density lipoprotein, total cholesterol, triglycerides, estimated glomerular filtration rate. Model 2 additionally adjusted for within-participant mean BMI across longitudinal measures. In stratified analyses, corresponding variable for the stratification was not included (e.g., sex was excluded in females or males only analysis). Stratified analysis among underweight group was not performed due to the small sample size and number of events. P-values indicate the interaction between BMI variability and the variables of stratification.

All stratified analysis results showed a consistent direction of hazardous effect of BMI variability (**Table 2 and Figure 3)**. In Model 2, the within-participant mean BMI was associated with greater risk of incident HF in all subgroups, except for the normal weight and overweight groups (**Supplementary Table 1)**. Stratified analysis among the underweight group was not performed due to the small sample size (N=445) and number of events (13 incident heart failure cases). We identified a suggestive interaction between BMI variability and sex (SD: p = 0.086; CV: p = 0.12), with potentially stronger associations observed among females than males in the fully adjusted models for both SD (female HR 1.07, 95% CI 1.03 - 1.11, p = 0.0009; male HR 1.03, 95% CI 1.00 - 1.06, p = 0.0674) and CV (female HR 1.09, 95% CI 1.04 - 1.14, p < 0.0001; male HR 1.04, 95% CI 1.01 - 1.08, p = 0.0178) (**Table 2 and Figure 3)**. Interactions with age, diabetes, hypertension, and BMI weight status category at enrollment were not identified.

## Discussion

This is the first large study that reports a positive association between BMI variability and incident HF, independent of known HF risk factors and mean BMI, after excluding prevalent cancer cases that are susceptible to weight fluctuations. The associations of BMI variability with incident HF were consistent across all BMI categories. It has been shown in recent studies, that obesity plays a potential causal role in HF incidence based on single measurement of BMI ^10,22,23^. However, longitudinal multiple measures of time-varying BMI contain additional information that can better characterize an individual’s underlying metabolic condition. The associations between BMI variability and incidence of cardiovascular diseases such as coronary heart disease, and mortality outcomes have been established ^14,24^, but the relationship between BMI variability and incident HF in the general population remains unclear. Our study addresses an important knowledge gap in HF susceptibility across several risk factor categories. Potential sex differences may indicate that the hazardous effect of major changes in body weight impacts women more than men in HF onset. BMI variability can be included as an informative measurement of HF risk to better assess populations susceptible to HF.

Prior studies have noted an association between body weight change and adverse cardiovascular outcomes, including myocardial infarction, stroke, and mortality among patients with type 2 diabetes ^25^ or hypertension ^26^. Further, existing evidence has shown that body weight variability is associated with cardiovascular disease risk among patients with type 2 diabetes as compared to non-diabetic controls ^27^. Even though HF shares similar risk profile with these CVD outcomes, no statistically significant difference was observed comparing risk factor categories including diabetes and hypertension.

It is worth noting that this new evidence should not override the well-established benefits of weight loss in obese populations to prevent cardiovascular diseases, including HF ^28^. Our definition of BMI variability was solely based on the primary care data, which did not denote intentional or unintentional weight loss. Intentional weight loss via dieting or exercise is more likely among healthy individuals and might be subject to weight regain, whereas unintentional weight loss might be associated with some chronic medical conditions such as depression and diabetes, which have been linked with cardiovascular diseases ^29,30^. Further studies are needed to separate these two mechanisms. Our study has several potential limitations. First, our current assessment of BMI variability did not differentiate the subtypes of weight change such as weight loss, gain or cycling. Future studies are warranted to investigate the trajectories in BMI changes and evaluate the differences in risk profile. Second, our definition of HF did not consider the reduced or preserved ejection fraction HF subtypes. Future studies should consider subgroups of HF cases and explore potentially different role of BMI variability. Third, this study was restricted to White participants due to available data. HF has a higher prevalence among other race or ethnicity groups such as African Americans and Hispanics ^2^, that similar investigation of BMI variability need to be expanded to. Future studies need to assess the generalizability of our findings towards diverse populations that vary in geographic regions, demographic, and socioeconomic status, with the goal of addressing the increasing global burden of obesity and heart failure. Finally, this study only used data from participants who had multiple BMI measurements in the primary care data. Multiple primary care visits indicate a propensity to more comorbidities, even though we excluded prevalent cancer and HF at enrollment.

## Conclusion

BMI fluctuation is an independent predictor of HF incidence with consistent associations across risk factor subgroups. Future studies focusing on the underlying mechanisms and drivers of longitudinal BMI change could lead to new insights of weight management strategies for HF prevention and intervention.

## Data Availability

The dataset used for this study is available in the UK Biobank public repository. This research has been conducted using the UK Biobank Resource under Application Number "34031".

## Acknowledgements

This research has been conducted using the UK Biobank Resource under Application Number “34031”.

